# IMPAIRED OLFACTORY FUNCTION IN SUBSTANCE USE DISORDER

**DOI:** 10.1101/2025.10.09.25337086

**Authors:** Clara U. Raithel, Garrick T. Sherman, David H. Epstein, Thorsten Kahnt

## Abstract

The sense of smell plays a key role in guiding motivated behavior, and olfactory function is impaired in clinical populations with dysfunctional approach-avoidance behavior, including major depressive and alcohol use disorder (AUD). However, whether olfactory impairments are also observed in individuals with substance use disorders (SUDs) other than AUD is currently unknown. This study aimed to evaluate the relationship between olfactory function and SUDs. We tested olfaction in 40 individuals with substance use disorders (SUDs) other than AUD using the Sniffin’ Sticks odor identification and olfactory threshold tests, versus 112 controls. Group differences were assessed with linear regression models, with diagnosis (SUD vs. controls) as a predictor, controlling for age, sex and smoking. Across a diverse range of substances used, individuals with SUDs had significantly lower *identification* scores than those in the control group. In contrast, olfactory *thresholds* did not differ significantly by diagnosis overall. However, exploratory analyses showed that men with SUDs had lower olfactory threshold scores (i.e., higher thresholds) than men in the control group, a difference that was absent in women. These results suggest that olfactory function is impaired in individuals with SUDs relative to controls. There are several plausible pathways by which differences in olfaction could be related to differences in hedonic processing, but longitudinal studies are needed to clarify the timing of olfactory impairment relative to substance use or SUD symptomatology.

## INTRODUCTION

Olfaction plays a key role in guiding motivated behavior. For instance, odors can trigger approach responses toward objects of positive hedonic value (e.g., one’s favorite food item), or they can prompt avoidance of aversive or harmful objects (e.g., gas or rotten foods). As such, odors are crucial for mediating adaptive behavior and decision making. Moreover, anosmia (i.e., the loss of smell) is reliably associated with several psychiatric problems, especially major depressive disorder (Croy & Hummel, 2017; Croy, Symmank, et al., 2014; Taalman et al., 2017), though there is no consensus on whether anosmia is a marker, cause, consequence, or some combination of cause and consequence.

Another psychiatric problem sometimes associated with anosmia is substance use disorder (SUD) (Lombion-Pouthier et al., 2006). An SUD can be defined as a disorder of decision making, characterized by compulsive behaviors directed toward drug use at the expense of alternative reward-guided behaviors and despite long-term negative consequences (Agarwal et al., 2021; Volkow et al., 2016). In line with this, individuals with an SUD sometimes have diminished affective responses to nondrug rewards (e.g., food, social interactions, etc.) and a corresponding lack of motivation to pursue such rewards (Hagele et al., 2015; Hyatt et al., 2012; Joyner et al., 2019; Konova et al., 2012). Notably, there is anatomical overlap between the brain circuits involved in olfaction and decision-making, including the ventral prefrontal lobe (e.g., orbitofrontal cortex), the ventral striatum (e.g., olfactory tubercle), and the medial temporal lobe (e.g., amygdala, entorhinal cortex, hippocampus) (Dikecligil & Gottfried, 2024; Gottfried, 2010; Koob & Volkow, 2016; Volkow et al., 2013; Zilverstand et al., 2018). This raises the intriguing question of whether the ability to perceive intrinsically rewarding stimuli, such as pleasant odorants, is diminished in individuals with SUDs, and whether such differences, if present, are due to impaired processing on an early sensory or higher cognitive level.

The relationship between SUDs and olfactory function remains largely unexplored for most substances but has been examined for tobacco and alcohol. The direct effects of tobacco smoke on olfactory function, reversible through abstinence, complicate interpretation of data from smokers (Ajmani et al., 2017; Etter et al., 2013; Frye et al., 1990), and evidence is inconclusive for olfactory function in individuals using alternative forms of nicotine, such as vaping, or heated tobacco products (Majchrzak et al., 2020; Sever et al., 2024; Ventoso, 2023). Individuals with alcohol use disorder (AUD) (Brion et al., 2015; Maurage, Callot, Chang, et al., 2011; Maurage, Callot, Philippot, et al., 2011; Shear et al., 1992), or those whose drinking patterns indicate risk for it (Agarwal et al., 2023; Agarwal et al., 2024), show olfactory impairments. Olfactory impairments in AUD are correlated with AUD severity. Individuals with risky drinking habits show impairments in identifying specific odorants (Agarwal et al., 2024). Moreover, individuals with alcohol dependence (by DSM-IV criteria) show impaired odor discrimination, and individuals with Korsakoff syndrome display decreases in both discrimination and identification (Brion et al., 2015). Interestingly, olfactory thresholds appear unaltered in both groups (Brion et al., 2015).

Only a few studies have investigated olfactory function in the context of other substance use or SUDs. These studies have examined opioid use disorder (Haghshenas Bilehsavar et al., 2022; Perl et al., 1997), cocaine use (Gordon et al., 1990), and cannabis use (Chao et al., 2021; Kao et al., 2022). Of note, these studies use a heterogeneous set of methods to measure olfactory function and are often restricted to small sample sizes – making the existing evidence difficult to interpret. As such, it remains unclear if and how olfactory function is impaired in individuals who use or are addicted to other substances. Investigating this relationship could reveal important insights regarding the pathophysiology of SUD and ultimately provide new avenues to treatment and improving the quality of life in this patient population.

In the current study, we analyzed an existing dataset including formal assessments of both SUDs and olfactory function. We compared individuals with SUDs to controls with respect to odor identification, olfactory thresholds, or both. Based on preliminary reports in the literature (Brion et al., 2015; Haghshenas Bilehsavar et al., 2022; Maurage, Callot, Chang, et al., 2011; Maurage, Callot, Philippot, et al., 2011; Perl et al., 1997) we hypothesized that individuals with an SUD would have impaired odor identification performance and, although less commonly reported, higher olfactory thresholds. We also conducted exploratory analyses to assess sex-specific associations between SUD and olfactory function.

## METHODS AND MATERIALS

### Dataset

We analyzed de-identified data from screening protocols at the Office of the Clinical Director of the National Institute on Drug Abuse Intramural Research Program (NIDA IRP). As part of these protocols, volunteers interested in participating in research studies at NIDA IRP undergo various screening assessments to determine study eligibility. Of note, these protocols enroll individuals with current or past SUD, as well as those who have never had an SUD. Individuals with SUDs may have undergone treatment at the time of enrollment or at some other timepoint in the past. To be enrolled in the screening protocols, individuals had to be 1) between 18 and 99 years old, 2) proficient to read, write and understand English, 3) willing to comply with screening procedures and 4) willing and able to sign a written informed consent form.

### Study Sample Selection

We used data from individuals with SUDs and those without who had completed either the Sniffin’ Sticks Identification Test (n=82) and/or the Sniffin’ Sticks Threshold Test (n=76) (Hummel et al., 1997), as part of the screening procedures.

### Substance Use Disorder status

Group membership (SUD or control group) was determined by either the Mini-International Neuropsychiatric Interview (MINI) (Sheehan et al., 1998) or the Structured Clinical Interview for DSM-5 (SCID) (First, 2016), each of which determines the presence or absence of an SUD in the past 12 months. Individuals with only an alcohol use disorder (AUD) were not included in the current analyses. However, n=2 individuals with SUDs also had an AUD (**Table 1** and **Fig. 1**) and were included in the SUD group.

**Table 1:**
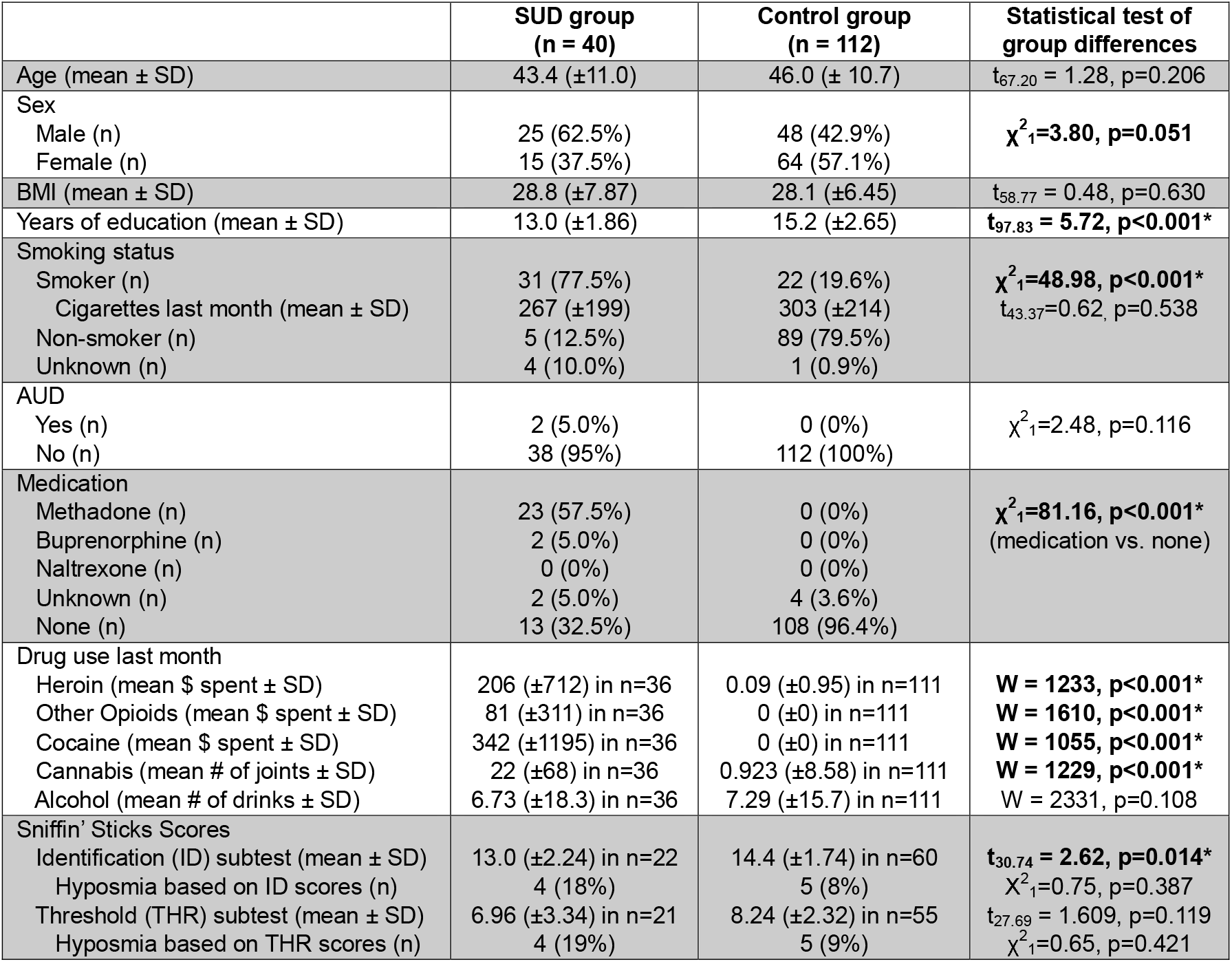
Demographic variables. * indicate p<0.05.

**Figure 1:**
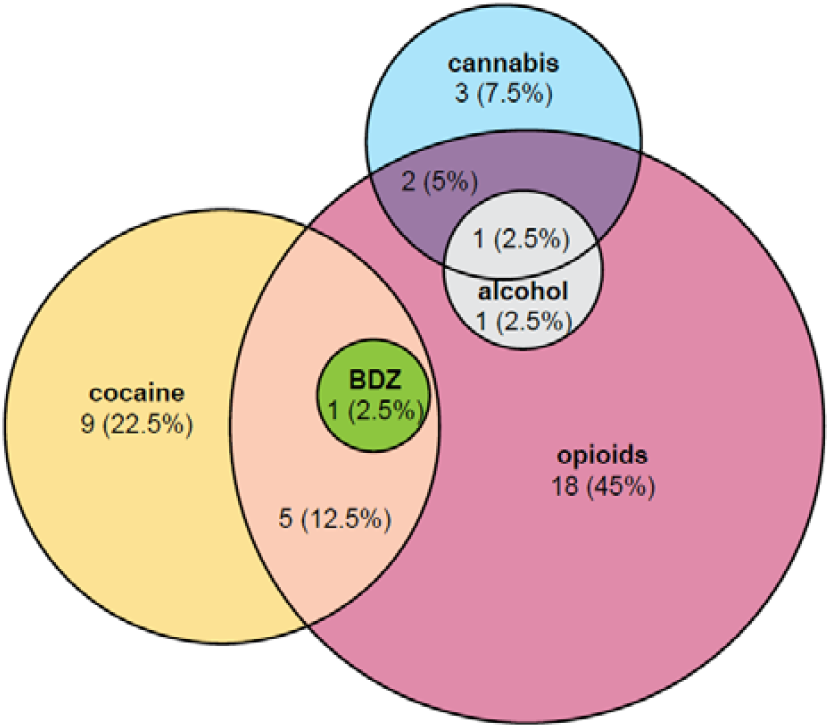
Overview of SUDs prevalent in the SUD group. Venn diagram illustrates the number and percentage (in parentheses) of participants in the SUD group with a given type of SUD. BDZ: benzodiazepine.

### Demographic variables

The following information was collected from participants: age, sex, Body Mass Index (BMI), smoking status, number of cigarettes smoked within the last 30 calendar days, and SUD-related medication. MINI data on the most problematic drug(s) and SCID data on differential diagnoses were combined to determine the type of SUDs prevalent in the SUD group (**Fig. 1**). Timeline Followback (Robinson et al., 2014; Roy et al., 2008; Sobell et al., 2003) data on drugs used within the last 30 calendar days were used to characterize drug use in both groups (**Table 1**).

### Assessments of olfactory function

#### Sniffin’ Sticks Identification Test

The Sniffin’ Sticks Identification Test was used to determine a participant’s ability to identify odorants. For this test, the participant was presented with one of 16 odorant-containing pens and asked to choose one out of four written answer options. The identification score corresponds to the number of correct responses and ranged between 0 and 16 points. The higher the score, the better the participant’s ability to identify odors.

#### Sniffin’ Sticks Threshold Test

The Sniffin’ Sticks Threshold Test was used to determine the minimum concentration at which an odorant can be perceived by the participant. On each trial, the participant was presented with three pens, only one of which contained an odorant. The other two contained solvent only. The participant’s task was to indicate which of the three pens contained the odorant. The test comprised a total of 16 pen triplets, with odorant in the target pen varying in absolute concentration. Starting with the lowest odorant concentration, a staircase paradigm was used to determine the lowest concentration a participant could reliably identify, with the final score ranging between 1 and 16 points. The higher the score, the lower the olfactory threshold, and the better the olfactory perceptual acuity of the participant. Of note, the dataset included one individual with a threshold score of 0, which is not possible given the described range of test results. We assume this was a mistake in recording and that this individual did not correctly identify any of the odorant containing pens. Accordingly, we manually changed this threshold score from 0 to 1. All statistical tests support the same conclusions regardless of whether this change is made.

A small number of participants completed either the olfactory identification test (n=2) or the olfactory threshold test (n=1) on multiple occasions (up to 3 times) at different time points. Data from these individuals were averaged across time points and this average score was then entered into regression models for analysis. In addition, a small number of participants completed both the olfactory identification test and the olfactory threshold test (n=6). Although both groups contained the same number of individuals completing both tests (n=3 each), these small sample sizes did not allow for meaningful statistical comparisons.

### Data analysis

To test associations between diagnosis and olfactory function, we used linear regression models, with SUD status as the main independent variable and olfactory function as the dependent measure. Age, sex and number of cigarettes smoked within the last 30 days were included as covariates because these variables are typically related to olfactory function; i.e., women (Sorokowski et al., 2019), younger individuals (Doty & Kamath, 2014; Oleszkiewicz et al., 2019), and nonsmokers (Ajmani et al., 2017; Frye et al., 1990) have a better sense of smell. Participants with missing data for any of these variables were excluded from the regression models (**Table 1**).

In a set of exploratory analyses following inspection of the data, we ran an additional regression model for olfactory identification scores using years of education as a covariate. Furthermore, the interaction term of sex and SUD status was included in the final model for olfactory thresholds as an independent measure, to account and test for sex-specific effects of SUD status on olfactory threshold scores.

T-tests, Wilcoxon Rank Sum tests, or Chi-square tests for independent samples were used to assess group differences in demographic variables (**Table 1**) and in the proportion of hyposmic individuals based on updated Sniffin’ Sticks normative data (Oleszkiewicz et al., 2019). All statistical tests were two-tailed with α=0.05. We used the eulerr package (**Fig. 1**) and ggstatsplot package (Patil, 2021) (**Figs. 2–3**) in R for data visualization.

**Figure 2:**
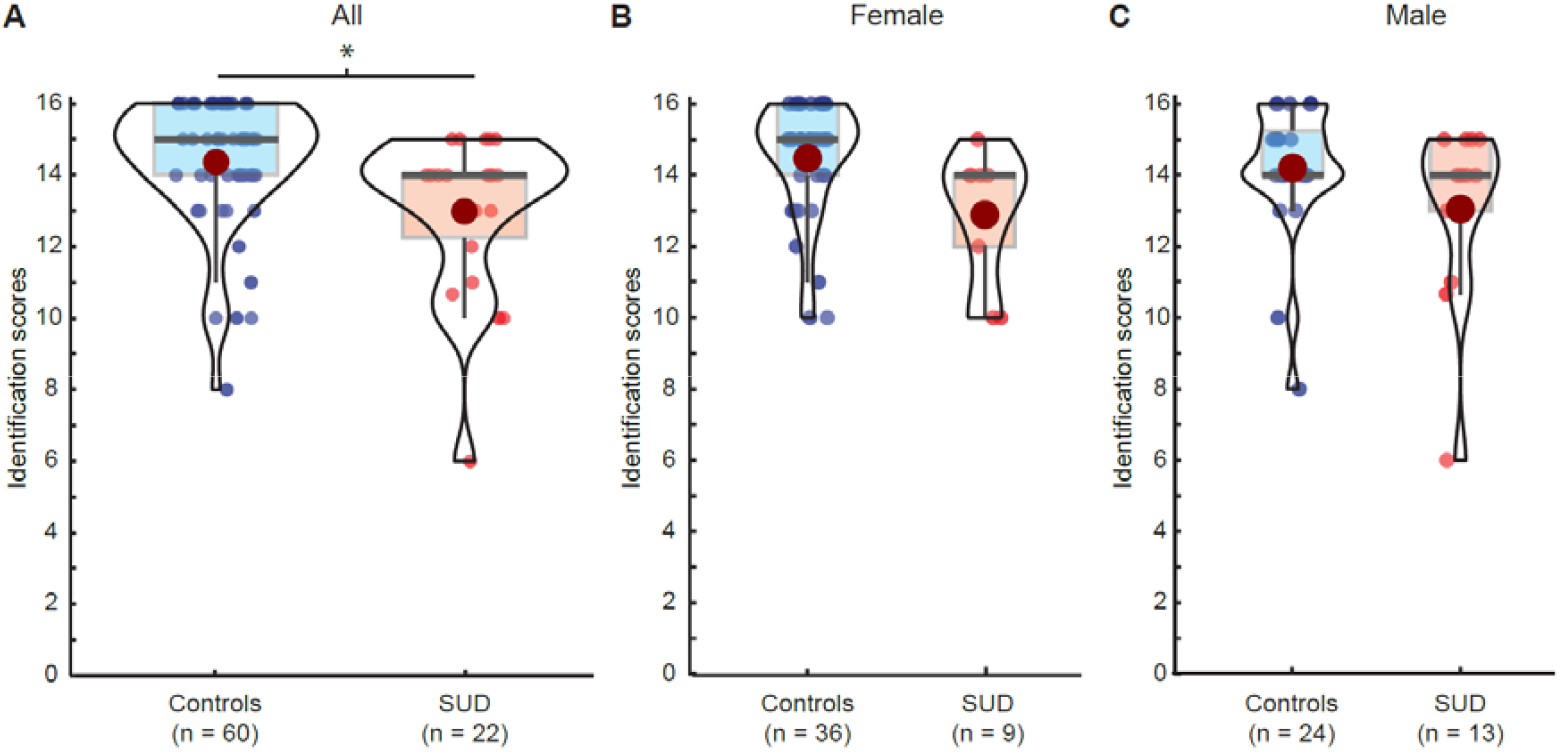
Odor identification scores in SUD and control groups. **A**. Olfactory identification scores in controls and individuals with an SUD. **B**. Olfactory identification scores in female controls and women with an SUD. **C**. Olfactory identification scores in male controls and men with an SUD. Red dots in violin plots indicate mean. * indicate p<0.05.

## RESULTS

### Demographic variables

As shown in **Table 1**, groups did not differ significantly in age (t_67.20_ = 1.28, p=0.206), BMI (t_58.71_ =0.48, p=0.630), or sex, although data for the latter indicated a trend towards a greater proportion of men in the SUD group (χ^2^_1_=3.80, p=0.051). The SUD group had fewer years of education (t_97.783_ =5.72, p<0.001) and a higher proportion of smokers (χ^2^_1_=48.98, p<0.001). Smokers in both groups had consumed a comparable number of cigarettes within the past 30 days (t_44.25_=0.60, p=0.552). Two individuals in the SUD group had a comorbid alcohol use disorder (AUD). Many individuals in the SUD group (n=25, 62.5%) were on medication for opioid use disorder.(Bell, 2014; Sullivan et al., 2013) Drug use measured using the Timeline Followback was significantly more prevalent in the SUD group for all substances assessed (heroin, other opioids, cocaine, cannabis) except alcohol (W=2331, p=0.104).

Opioid use disorder was the most common type of SUD in the SUD group (70%), with some participants having an opioid use disorder alone (45%) and some having comorbid cocaine use disorder (12.5%), cocaine and benzodiazepine use disorders (2.5%), cannabis use disorder (5%), cannabis and alcohol use disorders (2.5%), or alcohol use disorder (2.5%) (**Fig. 1**). 22.5% of the SUD group had cocaine use disorder only; 7.5% had cannabis use disorder only.

#### Lower odor identification scores in SUD

Odor identification scores were available for n=22 participants in the SUD group and n=60 participants in the control group. Scores were significantly lower in the SUD group (β_SUD_=−1.489, t_75_=−2.69, p=0.009; **Fig. 2**, and **Table 2**). The only other variable significantly associated with identification scores was age (β_age_=−0.056, t_75_=−2.17, p=0.033). The remaining variables had nonsignificant associations with identification, descriptively pointing in the expected directions (**Table 2**).

**Table 2:**
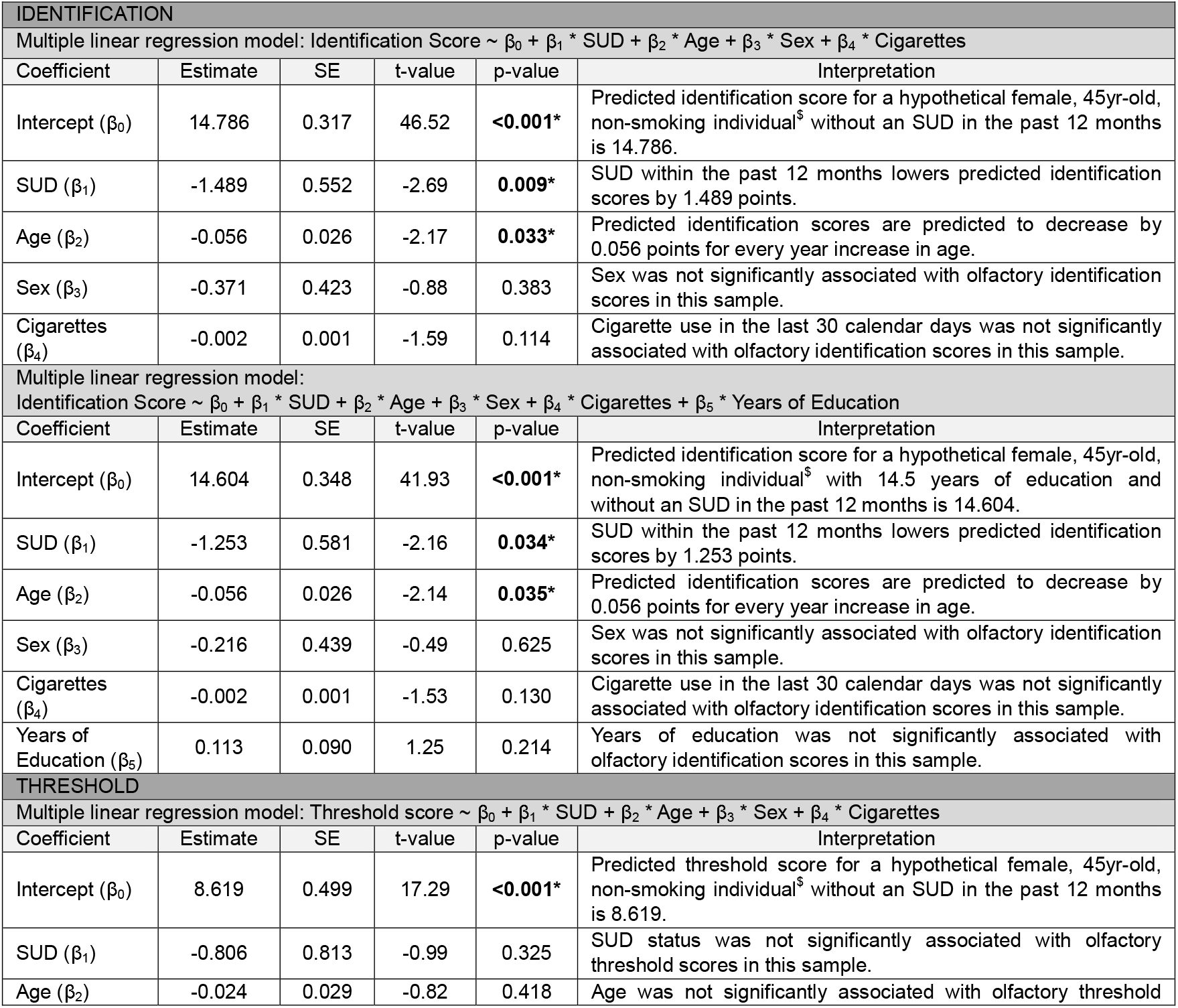
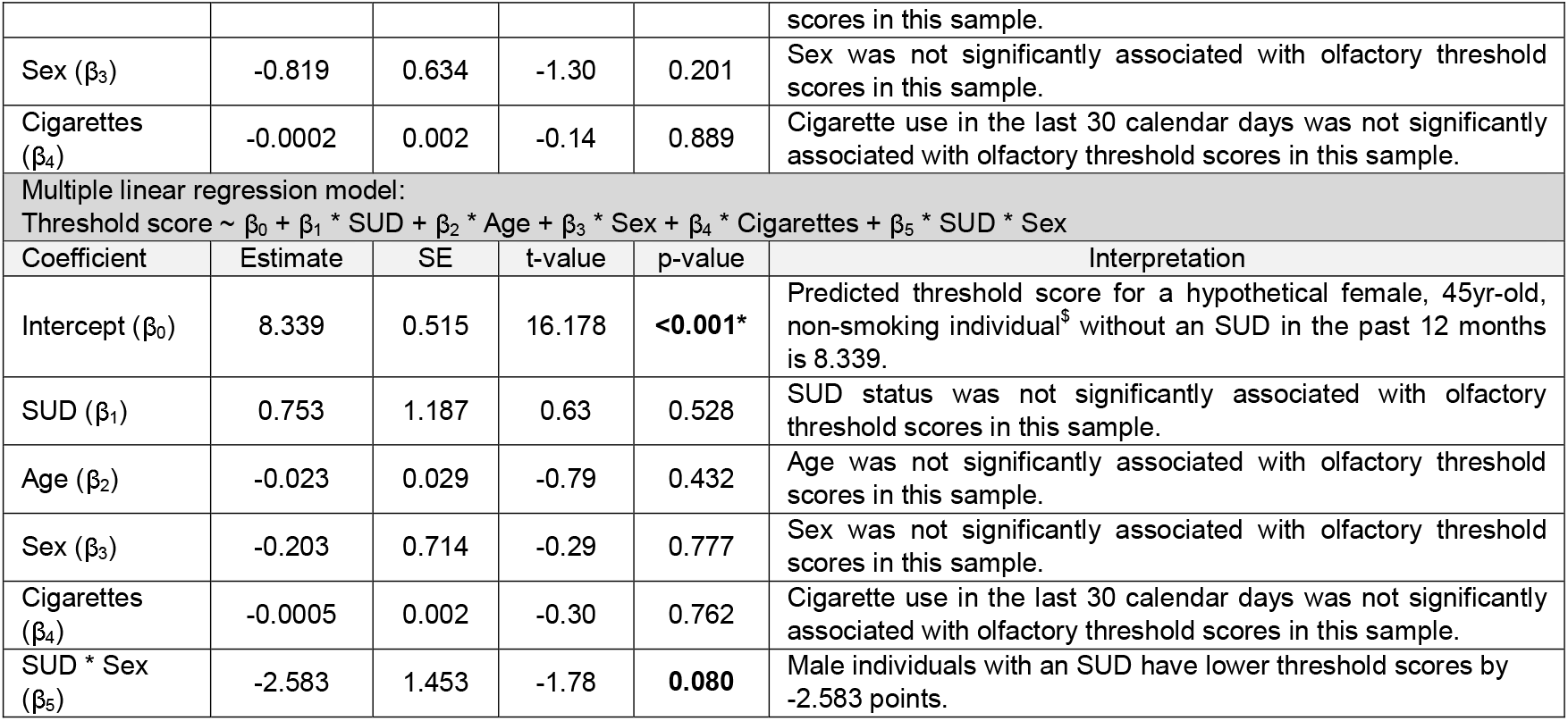
Results of Regression Models. Note that Age and Years of Education were mean centered before entering the regression model to allow for easier interpretation of βcoefficients. SE: standard error. $ please note these are interpretations of regression coefficients for a hypothetical individual and do not refer to an actual subject. * indicate p<0.05.

Given that groups differed in years of education (**Table 1**) and odor identification scores may be sensitive to educational attainment (Fornazieri et al., 2019; Gögele et al., 2024), we controlled for education in a second regression model. In this model, SUD status was still significantly associated with odor identification scores (β_SUD_=−1.253, t_74_=−2.16, p=0.034), suggesting that group differences in odor identification were not simply attributable to group differences in education.

Although the SUD group had a significantly lower mean score for olfactory identification, and a nominally greater proportion of scores indicating hyposmia (**Table 1**), individuals with SUDs were not more likely to be diagnosed as hyposmic based on these scores than individuals in the control group (χ^2^_1_=0.75, p=0.387).

### Olfactory thresholds are not significantly associated with SUD status

Olfactory threshold scores were available for n=21 participants in the SUD group and n=55 participants in the control group. Although the mean threshold in the SUD group was nominally worse (i.e., lower score, higher threshold scores) than in the control group (**Fig. 3A**), the group difference was not statistically significant (β_SUD_=−0.806, t_68_=−0.99, p=0.325; **Table 2**). Similarly, individuals with an SUD did not have a greater risk of being diagnosed as hyposmic based on these scores compared to individuals in the control group (χ^2^_1_=0.65, p=0.4212).

**Figure 3:**
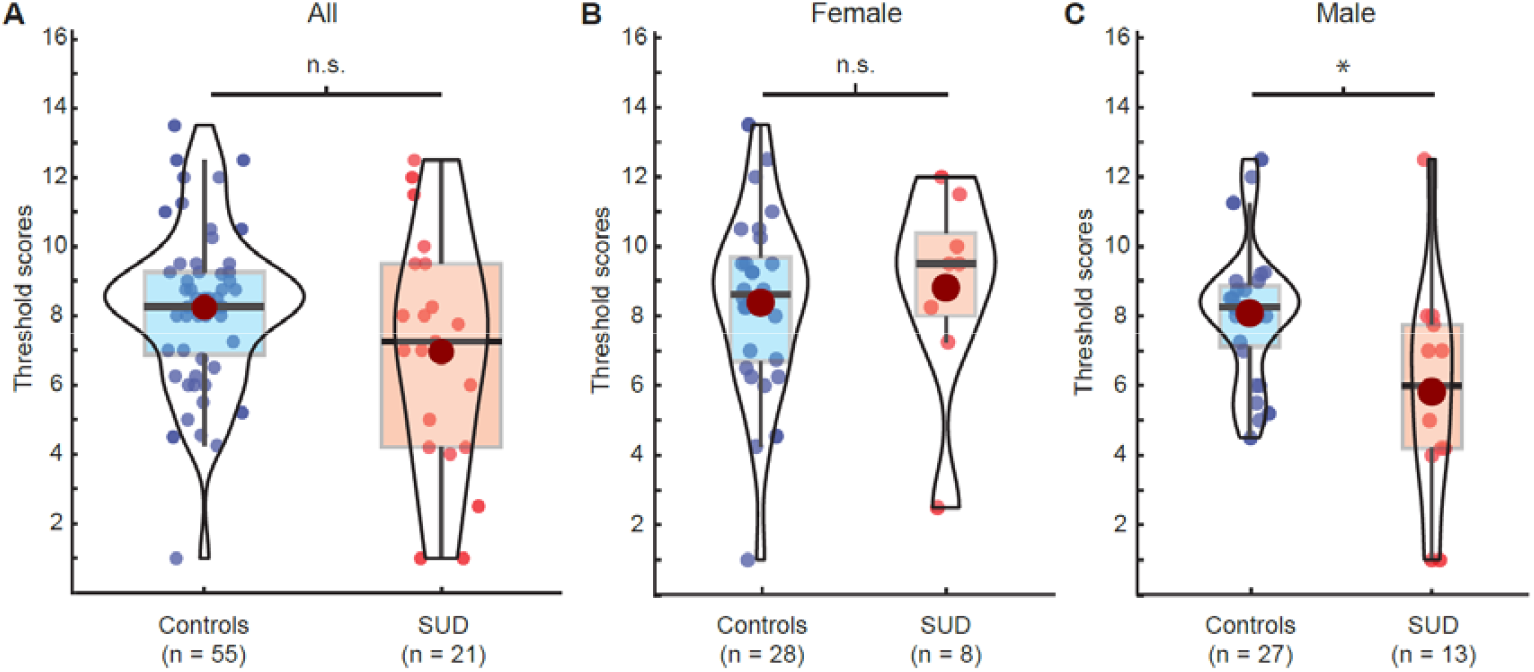
Olfactory threshold scores in SUD and control groups. Higher scores indicate lower thresholds (i.e., better performance). **A**. Olfactory threshold scores in controls and individuals with an SUD. **B**. Olfactory threshold scores in female controls and women with an SUD. **C**. Olfactory threshold scores in male controls and men with an SUD. Red dots in violin plots indicate mean. * indicate p<0.05.

Inspecting olfactory threshold scores for females and males separately, we found that women with an SUD tended to have nominally better scores than women in the control group, whereas men in the SUD group had nominally worse olfactory threshold scores than men in the control group (**Figs. 3B, 3C**). We followed up with a regression including the interaction between sex and SUD status. There was a trend-level effect for the interaction term (β_SUD*sex_=−2.583, t_67_=−1.78, p=0.080), suggesting that nominally worse olfactory threshold scores for the SUD group were driven by the men. Indeed, post-hoc simple regression models (controlling for age and smoking) performed separately for men and women, showed that the association in men was statistically significant in the predicted direction (β_SUD_=−2.352, t_34_=−2.47, p=0.019) but nominally in the opposite direction (though not statistically significant) in women (β_SUD_=2.129, t_31_=1.57, p=0.126).

## DISCUSSION

Olfaction plays a key role in many motivated behaviors that are disrupted in SUD. In this study, we have shown that individuals with an SUD exhibit deficits in odor identification compared to those without (even after controlling for smoking, age, and education), while olfactory thresholds were preserved. These results suggest that higher-order olfactory processing in the presence of an SUD is less efficient on average than in the absence of an SUD.

Our findings in individuals with SUDs involving opioids, cocaine, and cannabis parallel prior findings in individuals with AUD (Brion et al., 2015; Maurage, Callot, Chang, et al., 2011; Maurage, Callot, Philippot, et al., 2011). In both populations, identification (or discrimination) scores were lower compared to controls, while threshold scores were not. This pattern of results may indicate that olfactory function in SUDs is altered not at the peripheral level (i.e., the olfactory epithelium inside the nose), but instead at the level of central processing in the brain. Candidate regions include the ventral prefrontal lobe, as well as medial temporal lobe structures known to play a critical role in odor perception (Gottfried & Zald, 2005; Howard et al., 2015; Howard & Kahnt, 2017; Howard et al., 2009; Jiang et al., 2017; Jin et al., 2015; Kehl et al., 2024; Zelano et al., 2016; Zelano et al., 2011).

Despite preserved olfactory thresholds in individuals with SUDs overall, our exploratory analyses suggested a sex difference whereby olfactory thresholds were higher (i.e., worse) in men with SUDs compared to men in the control group. Formal tests of the corresponding interaction gave trend-level support (p=.080). Perhaps countervailingly, a recently published study of 20 men with heroin dependence (by DSM-IV criteria) showed lower discrimination and identification but preserved thresholds compared to 20 controls (Haghshenas Bilehsavar et al., 2022). Small sample sizes make it difficult to determine the extent to which that finding is discrepant from ours. Future work should account for possible effects of specific types of drugs and routes of administration (e.g., intranasal vs. intravenous), as they may differentially affect peripheral and/or central olfactory systems.

Observed differences in olfactory function between individuals with and without an SUD should be interpreted in relative, not in absolute terms. That is, individuals in the SUD group were not more likely to be diagnosed as hyposmic than the controls based on these scores. While group differences may be more pronounced in samples in which drug use (e.g., alcohol use, **Table 1**) among the control group is more tightly controlled, our findings do not rule out that the relative deficits observed in individuals with SUDs are potentially meaningful in their everyday lives, and hence clinically relevant. Assessments of qualify of life, as performed in studies on alcohol use (Agarwal et al., 2023), would provide valuable insight regarding this question.

Our results show a relationship between SUD and olfactory function, but it is a correlational one. We can think of at least four different models that could explain this relationship. First, as often assumed in the literature (Ackerman & Kasbekar, 1997; Agarwal, 2022; Heinbockel & Wang, 2015), chronic high-level exposure to drugs may cause olfactory changes, perhaps through neurotoxic effects on relevant brain areas. Second, changes specific *to SUDs* may alter olfactory function, because brain areas that undergo changes and adaptations in SUDs (Koob & Volkow, 2016; Volkow et al., 2013; Zilverstand et al., 2018) partially overlap with those involved in olfactory processing (Gottfried & Zald, 2005; Howard et al., 2015; Howard & Kahnt, 2017; Jin et al., 2015; Zelano et al., 2016; Zhou et al., 2019). Third, as has occasionally been proposed (Maurage, Callot, Philippot, et al., 2011) olfactory problems may precede and contribute to the development of an SUD: a dysfunctional sense of smell may impair the ability to derive hedonic value from nondrug rewards in everyday life (such as food, social interactions, etc.), biasing individuals to seek out other rewarding stimuli, such as drugs, ultimately predisposing them to an SUD. Lastly, it is also possible that preexisting alterations in brain areas involved in decision making and olfaction (e.g., olfactory tubercule in the ventral striatum, amygdala, orbitofrontal cortex, etc.), may independently predispose individuals to both SUDs and olfactory impairments. Studies with adequate experimental designs (e.g., longitudinal studies, causal manipulations, etc.) are needed to test these different models.

While it may take time to resolve the question surrounding the causal mechanisms underlying the association between SUD and olfactory function, existing evidence suggests that olfactory testing could be an important tool in the diagnosis of SUD, as well as the development of effective treatment strategies. This is especially important as impairments in olfactory function are associated with lower quality of life (Croy, Nordin, et al., 2014; Zou et al., 2021), and can cause or exacerbate nutritional deficiencies (Aschenbrenner et al., 2008; Gopinath et al., 2016). Both factors are relevant for SUD populations, whose members often report lower quality of life and struggle with malnutrition, even (and perhaps specifically) during extended periods of abstinence (Jeynes & Gibson, 2017; Parvin et al., 2024). Perhaps olfactory training, commonly used in other clinical populations (Hummel et al., 2009; Konstantinidis et al., 2013; Pieniak et al., 2022; Yaylaci et al., 2023), can help to (at least partly) restore olfactory function in SUD patients and increase their quality of life and facilitate abstinence and recovery.

Our study bears several limitations. Olfactory identification and olfactory threshold scores were available from the same individuals in only 6 cases (n=3 per group), preventing us to test the direct association between olfactory identification and olfactory thresholds in individuals with SUDs. Furthermore, drug use in the control group was not strictly controlled for, and alcohol consumption in the past 30 days was similar between the groups (**Table 1**). However, on the plus side, this means that one might expect even stronger group differences in olfactory function if these variables are better controlled, and that the differences in olfactory function observed here are not confounded by differences in alcohol intake.

Taken together, our findings highlight olfactory impairments in individuals with an SUD and emphasize the need for a more mechanistic understanding of this relationship. Investigating the causal link will not only advance our basic scientific knowledge but may ultimately provide an opportunity for the development of preventative measures, early diagnostic tools, as well as treatment strategies.

## ACKNOWLEDGMENTS

The authors would like to thank the clinical and research staff involved in patient care, data collection, and other support, in the NIDA IRP Office of the Clinical Director.

## AUTHOR CONTRIBUTIONS

Clara U. Raithel: Conceptualization, Methodology, Formal analysis, Data Curation, Writing – Original Draft, Writing – Review & Editing, Visualization; Garrick Sherman: Conceptualization, Data Curation, Writing – Review & Editing; David H. Epstein: Conceptualization, Writing – Review & Editing; Thorsten Kahnt: Conceptualization, Methodology, Resources, Writing: Review & Editing, Supervision, Funding Acquisition.

## ETHICS DECLARATIONS

### Ethics approval

This study used de-identified data that were initially collected with informed consent under a screening protocol of the NIDA IRP OCD, which was approved by the Institutional Review Board of the NIH.

## DATA AVAILABILITY

The de-identified data set will be made available on a publicly available repository upon acceptance of the manuscript for publication.

